# Patterns and correlates of alcohol and other substance use among adults in two communities in Cape Town: a cross-sectional study

**DOI:** 10.64898/2026.10.12.26365289

**Authors:** Andreas D. Haas, Mpho Tlali, John Joska, Leigh L. van den Heuvel, Stephan Rabie, Noleen Williams, Onke Maniwe, Nur Khan, Lucia Madolo, Chido Chinogurei, Confidence Mothiba, Timothy Mountford, Soraya Seedat, Catherine Orrell, Kristina Adorjan, Mary-Ann Davies, Leigh Johnson, Reshma Kassanjee

**Affiliations:** Department of Psychiatry and Psychotherapy, University of Bern, Switzerland; Institute of Social and Preventive Medicine, University of Bern, Switzerland; Centre for Infectious Disease Epidemiology & Research, School of Public Health, University of Cape Town, South Africa; HIV Mental Health Research Unit, Neuroscience Institute, Department of Psychiatry and Mental Health, University of Cape Town, South Africa; Department of Psychiatry, Faculty of Medicine and Health Sciences, Stellenbosch University, Cape Town, South Africa; Graduate School for Health Sciences, University of Bern, Switzerland; Provincial Health Data Centre, Western Cape Department of Health and Wellness, Cape Town, South Africa; Desmond Tutu HIV Centre, Institute of Infectious Disease and Molecular Medicine, University of Cape Town, South Africa; HIV and Other Infectious Diseases Unit, South African Medical Research Council, South Africa; Institute of Psychiatric Phenomics and Genomics, LMU University Hospital, LMU Munich, Munich, Germany

## Abstract

**Background:** We aimed to characterize substance use patterns, and their correlates, among adults with and without HIV at two healthcare facilities in Cape Town, South Africa.

**Methods:** We recruited adults with and without HIV from two primary healthcare facilities. We assessed alcohol use with the AUDIT-C and other substance use with the WHO-ASSIST questionnaires. We classified participants with no lifetime drug use and no or low-risk alcohol use as “low risk” and fitted latent class models among remaining participants to identify additional substance use patterns. We used multinomial logistic regression to examine factors associated with substance use patterns.

**Results:** Among 602 participants, 265 (44.0%) were “low risk”. Latent class analysis among the remaining 337 identified three patterns: “higher-risk alcohol use” (N=168; 27.9%), “cannabis use” (universal cannabis use with mixed alcohol risk; N=114; 18.9%), and “mixed illicit drug use” (predominantly no or low-risk alcohol use with use of multiple drugs; N=55; 9.1%). Participants younger than 45 years were less likely to be in the low-risk group. Male sex was associated with lower probability of the low-risk group and higher probability of the higher-risk alcohol use and cannabis use classes. HIV was associated with higher probability of higher-risk alcohol use. One facility had a substantially higher probability of mixed illicit drug use.

**Conclusions:** We identified distinct higher-risk substance use profiles that varied strongly by age, sex, and facility. These findings support integrating alcohol and substance use screening and management into primary care and tailoring responses to local substance use patterns.

## 1. Introduction

In South Africa, harmful alcohol and illicit drug use are common and have increased over time. National survey data estimated risky, hazardous, or harmful alcohol use at 7.4% in 2005 and 10.3% in 2017.^1,2^ The prevalence of past-3-month illicit drug use increased from 1.5% in 2002 to 10.0% in 2017, reflecting increases in cannabis, opioid, cocaine, and amphetamine use.^3^ These national trends obscure important local and subgroup-specific variation. For example, in the Western Cape, methamphetamine became widely used especially among adolescents, reflected in a surge in treatment admissions in this age group in the mid-2000s,^4,5^ while national prevalence estimates in the general population remained relatively low at 1.5% in 2017.^3^

The same South African national survey data also identified important demographic, geographic, socioeconomic, and psychosocial correlates of harmful alcohol and drug use. Harmful alcohol use is more common among males and in urban areas.^1,2^ Drug use has been found to be associated with male sex, younger age, urban areas, unemployment, intimate partner violence, psychological distress, and harmful alcohol use.^3^

Most South African studies have examined individual substances separately, providing limited insight into patterns of concurrent use.^1–3^ Available evidence suggests that polysubstance use is common and frequently involves combinations of methamphetamine, cannabis, sedatives like methaqualone, cocaine or crack cocaine, and opioids, including heroin and heroin-containing mixtures, such as nyaope or whoonga.^6–8^ However, evidence on distinct patterns of concurrent substance use in South Africa remains limited to selected populations of people who use drugs, and female sex workers living with HIV.^6–8^

People with HIV represent an important population in the study of substance use in South Africa, given the country’s high HIV prevalence and the additional burden of psychosocial stress,^9,10^ HIV-related stigma,^11^ and mental health problems experienced by many people with HIV.^12,13^ Furthermore, people with HIV require lifelong adherence to antiretroviral therapy to prevent HIV-related morbidity and mortality, and alcohol and drug use have been associated with poorer medication adherence,^12^ viral non-suppression,^12^ disengagement from HIV care^14^, and increased mortality.^14,15^ In addition, HIV, mental health problems, and substance use are shaped by overlapping social and structural determinants, including poverty, unemployment, and violence.^16^

Despite the high prevalence of alcohol and other substance use in South Africa, important gaps remain in understanding distinct patterns of concurrent alcohol and substance use and how these vary by age, sex, and across socioeconomically disadvantaged communities, where community-specific substance use patterns may translate into differing service needs. Among people with HIV, characterizing substance use patterns may also help inform the recommended integration of screening and treatment services into HIV care.^17^

To address these gaps, we conducted a cross-sectional study among adults with and without HIV attending two primary healthcare facilities in socioeconomically disadvantaged communities in Cape Town. We assessed alcohol and other substance use using validated screening tools and used latent class analysis to identify distinct patterns of use. We then examined how these patterns varied by age, sex, study facility, and HIV status and assessed their clinical relevance against psychiatric diagnoses of alcohol and substance use disorders.

## 2. Methods

### Study design and participants

We conducted a cross-sectional study of adults, aged 18 years or older, who spoke English, Xhosa, or Afrikaans, and provided written informed consent, recruited between March 2023 and September 2024 from two primary care facilities in Cape Town, South Africa. Participants were interviewed in their preferred language (English, Xhosa, or Afrikaans). We used quota sampling to balance recruitment by sex and age group (18–29, 30–44, ≥45 years) and targeted a 2:1 ratio of participants with and without HIV. Most participants with HIV were recruited from antiretroviral therapy patient steams and those without HIV from outpatient services during routine or acute care visits.

### Setting

The two primary care facilities serve low-income communities established through forced removals under apartheid and now characterized by persistent socioeconomic disadvantage, high unemployment, gang violence, drug use, and limited access to mental health care, although the communities differ in sociodemographic composition, housing conditions, and HIV burden.^18^ The community served by Facility 1 has more informal housing, poorer household infrastructure, and higher HIV prevalence, whereas gang violence is particularly salient in the community served by Facility 2.

### Measures

Research assistants collected data on mental health symptoms, substance use, and sociodemographic, behavioral, and medical characteristics using screening instruments and structured questionnaires. Trained research nurses administered the MINI International Neuropsychiatric Interview (MINI 6.0)^19^ to establish diagnoses according to the Diagnostic and Statistical Manual of Mental Disorders, Fourth Edition (DSM-IV), including past year alcohol use disorder, past year other substance use disorders, and a broad range of mental disorders.

We assessed past-year alcohol use with the 3-item Alcohol Use Disorders Identification Test-Concise (AUDIT-C),^20^ which measures self-reported drinking frequency, typical consumption, and heavy episodic drinking (score range 0–12). We classified scores as low risk (0–3 males, 0–2 females), moderate risk (4–7 males, 3–7 females), or high risk (≥8).

We assessed cannabis, amphetamine, cocaine, sedative, hallucinogen, and opioid use with the Alcohol, Smoking and Substance Involvement Screening Test (ASSIST).^21^ The ASSIST assesses lifetime use and, among ever-users, past-3-month use, craving, health or social problems, role impairment, and unsuccessful attempts to cut down, generating substance-specific risk scores (0–39). For latent class analysis, we categorized cannabis use as no lifetime, low risk (0–3), or moderate-to-high risk (≥4), amphetamine use as no versus any lifetime use, and combined cocaine, sedative, hallucinogen, and opioid use into a binary indicator of any lifetime “other substance” use because these substances were uncommon.

HIV status was ascertained from clinic records or, when recent documentation was unavailable, by serial rapid testing according to the South African public-sector algorithm. Participants with two reactive tests were classified as HIV positive and referred for HIV care, while those with a nonreactive screening test were classified as HIV negative. Further details on the testing procedures have been reported previously.^22^

We assessed sociodemographic characteristics including age, sex, employment status in the previous 12 months (no employment, past but not current employment, or current employment), educational attainment (no formal education, primary education, or secondary or tertiary education), and marital status (single, married or cohabiting, or widowed, separated, or divorced).

### Statistical analysis

We used descriptive statistics to summarize participants’ sociodemographic and clinical characteristics by study facility.

We identified substance use patterns using a two-stage approach to address the high prevalence of non-use and the resulting zero inflation and sparse higher-risk categories in some substance use indicators. First, we classified participants with no lifetime drug use and no or low-risk alcohol use (AUDIT-C score, ≤2 for females and ≤3 for males) as an a priori low-risk group. Second, among the remaining participants, we fit latent class models using AUDIT-C risk category (low, moderate, or high), cannabis use risk (no lifetime use, low-risk use, or moderate- to high-risk use), lifetime amphetamine use, and lifetime other substance use. We fit latent class models with 2 to 5 classes using generalized structural equation modeling with ordinal logistic models. We compared model fit across candidate models using the Akaike information criterion (AIC) and Bayesian information criterion (BIC). We selected the final model primarily based on the BIC, with consideration of the AIC, class size, and interpretability. After model selection, we estimated posterior probabilities of class membership for each participant and assigned participants to the class with the highest posterior probabilities. We assessed class separation using entropy. Missing data on the alcohol and substance use indicators used to define the latent classes were handled within the maximum-likelihood estimation, allowing participants with partially observed indicators to contribute information from their observed responses.

To assess the construct validity and clinical relevance of the latent substance use classes, we examined whether class membership was associated with independent MINI diagnoses of alcohol use disorder and substance use disorder using multinomial logistic regression. We estimated adjusted relative risk ratios (RRRs) with 95% confidence intervals (CIs), adjusting for age group, sex, HIV status, and facility.

To quantify associations between latent substance use classes and age group, sex, HIV status, and facility, we estimated adjusted RRR with 95% CIs using multinomial logistic regression with the low-risk class as the reference category. To aid interpretation, we estimated adjusted predicted probabilities and average marginal effects (AME) from the fitted multinomial model, averaging over the observed distribution of the remaining covariates. AMEs were expressed as adjusted percentage-point differences in the predicted probability of each class for each level of age group, sex, HIV status, and facility. We present AME as the primary measure because they are directly interpretable on the absolute probability scale and illustrate how covariates shift the probability of class membership. Adjusted predicted probabilities are presented as complementary measures of absolute class probability.

We used complete-case data for multinomial logistic regression analyses because missing data were minimal. Statistical analyses were conducted in Stata version 18 (StataCorp LLC, College Station, TX, USA) and R version 4.4.2 (R Foundation for Statistical Computing, Vienna, Austria).

### Ethical considerations

The study received ethical approval from the Human Research Ethics Committee of the University of Cape Town (reference 129/2022) and the Cantonal Ethics Committee of Bern, Switzerland (reference 150/14). All participants provided written informed consent. In accordance with guidance from the University of Cape Town Human Research Ethics Committee, we do not report facility names, exact locations, or detailed contextual characteristics, such as population group and interview language, to minimize the risk of stigmatizing communities.

## 3. Results

### Characteristics of the study population

Of the 602 participants included in the analysis, 401 (66.6%) were recruited at Facility 1 and 201 (33.4%) at Facility 2 (Table 1). Overall, 394 participants (65.4%) were living with HIV, 386 (64.1%) were female, and the median age was 39 years (IQR, 27-48). In both facilities, slightly more than half of participants had primary education, and about two-thirds had not been employed during the previous 12 months.

**Table 1.** Participant sociodemographic and clinical characteristics by study site.

|  | Facility 1<br>N=401 (66.6%) | Facility 2<br>N=201 (33.4%) | Total<br>N=602 (100.0%) |
| --- | --- | --- | --- |
| HIV status |  |  |  |
| Positive | 318 (79.3%) | 76 (37.8%) | 394 (65.4%) |
| Negative | 82 (20.4%) | 124 (61.7%) | 206 (34.2%) |
| Missing | 1 (0.2%) | 1 (0.5%) | 2 (0.3%) |
| Age, years |  |  |  |
| 18-29 | 131 (32.7%) | 45 (22.4%) | 176 (29.2%) |
| 30-44 | 122 (30.4%) | 100 (49.8%) | 222 (36.9%) |
| 45+ | 148 (36.9%) | 56 (27.9%) | 204 (33.9%) |
| Median (IQR) | 39 (25-48) | 38 (30-46) | 39 (27-48) |
| Sex |  |  |  |
| Female | 252 (62.8%) | 134 (66.7%) | 386 (64.1%) |
| Male | 149 (37.2%) | 67 (33.3%) | 216 (35.9%) |
| Highest level of Education |  |  |  |
| No education | 28 (7.0%) | 26 (12.9%) | 54 (9.0%) |
| Primary | 224 (55.9%) | 119 (59.2%) | 343 (57.0%) |
| Secondary/Tertiary | 148 (36.9%) | 55 (27.4%) | 203 (33.7%) |
| Missing | 1 (0.2%) | 1 (0.5%) | 2 (0.3%) |
| Employment |  |  |  |
| None (past 12m) | 279 (69.6%) | 118 (58.7%) | 397 (65.9%) |
| Past 12m, not current | 0 (0.0%) | 48 (23.9%) | 48 (8.0%) |
| Current | 118 (29.4%) | 33 (16.4%) | 151 (25.1%) |
| Missing | 4 (1.0%) | 2 (1.0%) | 6 (1.0%) |
| Marital status |  |  |  |
| Single | 278 (69.3%) | 98 (48.8%) | 376 (62.5%) |
| Married/living together | 80 (20.0%) | 67 (33.3%) | 147 (24.4%) |
| Widowed/separated/divorced | 43 (10.7%) | 35 (17.4%) | 78 (13.0%) |
| Missing | 0 (0.0%) | 1 (0.5%) | 1 (0.2%) |
| Any mental disorder, current | 57 (14.2%) | 70 (34.8%) | 127 (21.1%) |
| Psychotic disorder | 13 (3.2%) | 1 (0.5%) | 14 (2.3%) |
| Bipolar-related disorders | 4 (1.0%) | 3 (1.5%) | 7 (1.2%) |
| Major depressive disorder | 30 (7.5%) | 42 (20.9%) | 72 (12.0%) |
| Generalized anxiety disorder | 3 (0.7%) | 39 (19.4%) | 42 (7.0%) |
| Post-traumatic stress disorder | 12 (3.0%) | 37 (18.4%) | 49 (8.1%) |
| Other anxiety disorders | 10 (2.5%) | 20 (10.0%) | 30 (5.0%) |
| Any substance use disorder, current | 125 (31.2%) | 69 (34.3%) | 194 (32.2%) |
| Alcohol use disorder | 119 (29.7%) | 23 (11.4%) | 142 (23.6%) |
| Substance use disorder | 12 (3.0%) | 56 (27.9%) | 68 (11.3%) |

Data are n (%) unless otherwise stated. Current mental disorders were defined based on Mini-International Neuropsychiatric Interview diagnoses. IQR=interquartile range.

Current mental disorders, as defined by the MINI, were more than twice as common at Facility 2 as at Facility 1 (34.8% vs 14.2%). Facility 2 had higher prevalences of major depressive disorder, anxiety, and post-traumatic stress disorders, whereas psychotic disorders were more prevalent at Facility 1 (Table 1).

Any current substance use disorders, defined based on the MINI as either alcohol use disorder or other substance use disorders, were common at both facilities and were diagnosed in about one-third of participants at each site. The pattern differed by type of substance. Alcohol use disorder was more prevalent at Facility 1, whereas non-alcohol substance use disorder was considerably more common at Facility 2 (Table 1).

### Alcohol use and substance use risk

Table 2 shows alcohol use risk based on the AUDIT-C and substance-specific lifetime use and ASSIST scores by study site. Alcohol use risk categories were broadly similar across facilities. Most participants were in the low-risk category (58.9% at Facility 1 and 64.7% at Facility 2), while 31.2% at Facility 1 and 23.9% at Facility 2 were in the moderate-risk category. High-risk alcohol use was identified in 6.5% of participants at Facility 1 and 10.4% at Facility 2.

**Table 2.** Alcohol use risk categories and substance-specific lifetime use by study site.

|  | Facility 1<br>N=401 (66.6%) | Facility 2<br>N=201 (33.4%) | Total<br>N=602 (100.0%) |
| --- | --- | --- | --- |
| Alcohol use risk (AUDIT-C) |  |  |  |
| Low | 236 (58.9%) | 130 (64.7%) | 366 (60.8%) |
| Moderate | 125 (31.2%) | 48 (23.9%) | 173 (28.7%) |
| High | 26 (6.5%) | 21 (10.4%) | 47 (7.8%) |
| Missing | 14 (3.5%) | 2 (1.0%) | 16 (2.7%) |
| AUDIT-C score, median (IQR) | 2 (0-4) | 0 (0-5) | 2 (0-4) |
| Cannabis use |  |  |  |
| Any lifetime use, n (%) | 81 (20.2%) | 46 (23.5%) | 127 (21.3%) |
| ASSIST score among lifetime users, median (IQR) | 4 (0-15) | 18 (12-20) | 10 (0-18) |
| Amphetamine use |  |  |  |
| Any lifetime use, n (%) | 3 (0.8%) | 46 (24.7%) | 49 (8.4%) |
| ASSIST score among lifetime users, median (IQR) | 10 (0-21) | 10 (6-30) | 10 (6-29) |
| Other substance use |  |  |  |
| Any lifetime use, n (%) | 4 (1.0%) | 42 (22.7%) | 46 (7.9%) |
| ASSIST score among lifetime users, median (IQR) | 12 (0-24) | 23 (6-33) | 23 (6-33) |
| Sedatives use |  |  |  |
| Any lifetime use, n (%) | 0 (0.0%) | 25 (13.7%) | 25 (4.3%) |
| ASSIST score among lifetime users, median (IQR) | NA | 26 (6-31) | 26 (6-31) |
| Cocaine use |  |  |  |
| Any lifetime use, n (%) | 4 (1.0%) | 6 (3.3%) | 10 (1.7%) |
| ASSIST score among lifetime users, median (IQR) | 12 (0-24) | 6 (3-6) | 6 (2-24) |
| Hallucinogens use |  |  |  |
| Any lifetime use, n (%) | 0 (0.0%) | 6 (3.3%) | 6 (1.0%) |
| ASSIST score among lifetime users, median (IQR) | NA | 3 (0-6) | 3 (0-6) |
| Opioids use |  |  |  |
| Any lifetime use, n (%) | 0 (0.0%) | 13 (7.2%) | 13 (2.3%) |
| ASSIST score among lifetime users, median (IQR) | NA | 30 (6-34) | 30 (6-34) |

Data are n (%) or median (IQR). Percentages are calculated among participants with available data for the respective measure and may therefore not use the full study-site denominator. Numbers of complete and missing observations are reported in Table S2. ASSIST scores are shown among participants reporting lifetime use of the respective substance. In the latent class analysis, other substance use was included as a composite category comprising sedatives, cocaine, hallucinogens, and opioids. These substances are also shown separately in this table. AUDIT-C=Alcohol Use Disorders Identification Test-Concise. ASSIST=Alcohol, Smoking and Substance Involvement Screening Test. IQR=interquartile range. NA=not applicable.

Lifetime cannabis use was similar across facilities (20.2% at Facility 1 and 23.5% at Facility 2), but median ASSIST scores among participants reporting lifetime cannabis use were higher at Facility 2 (18 vs 4). In contrast, lifetime amphetamine use and use of substances included in the other-substance category were uncommon at Facility 1 but substantially more frequent at Facility 2. Among participants reporting lifetime other-substance use, median ASSIST scores were also higher at Facility 2.

### Substance use classes

Among 602 participants, 265 (44.0%) were classified a priori into the low-risk class. Latent class analysis was conducted among the remaining 337 participants (56.0%). AIC and BIC values for models with 2 to 5 classes are shown in Table S1. The 3-class model with the lowest AIC and BIC was selected. Entropy for the selected model was 0.839.

The final classification comprised four substance use classes, including one low-risk class defined a priori and three classes identified by latent class analysis: low risk (n=265 [44.0%]), higher-risk alcohol use (n=168 [27.9%]), cannabis use (n=114 [18.9%]), and mixed illicit drug use (n=55 [9.1%]).

Figure 1 shows alcohol and other substance use risk categories by class. In the low-risk class, 65.3% reported no past-year alcohol use and 34.7% low-risk alcohol use. The higher-risk alcohol use class was characterized by universal moderate- to high-risk alcohol use with little other substance use. The cannabis use class was characterized by universal cannabis use, predominantly at low to moderate risk, with alcohol use at varying risk levels in most participants (83.3%) and little use of other substances. The mixed illicit drug use class was characterized by use of multiple substances, particularly amphetamines (79.2%), sedatives (44.0%), opioids (23.4%), cannabis (21.6%), hallucinogens (10.2%), and cocaine (6.0%). Alcohol use was less common in this class (27.3%). For all substances except alcohol and hallucinogens, use was predominantly at moderate or high risk. (Figure 1, Table S2).

**Figure 1.**
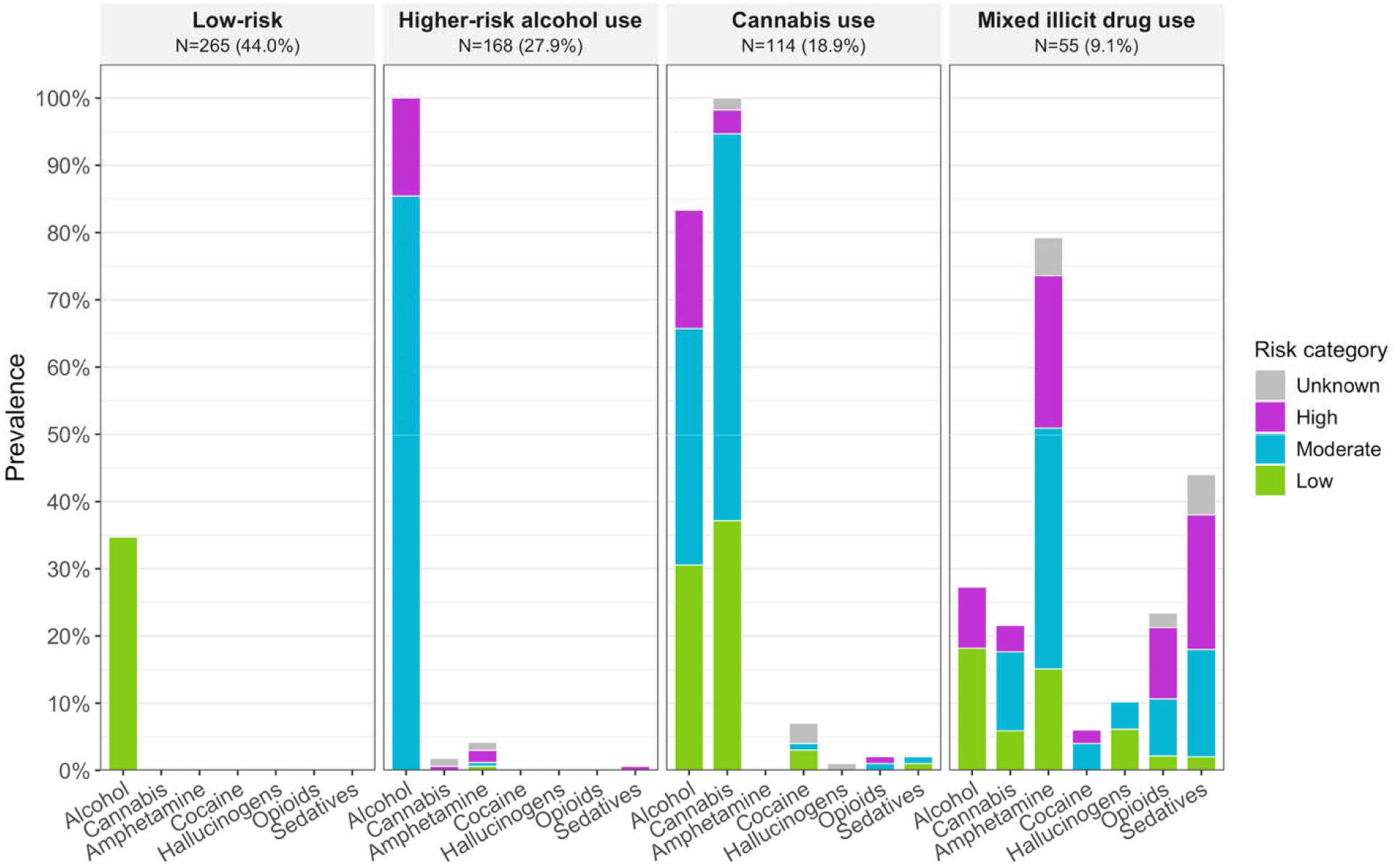
Prevalence of alcohol and substance use by risk category within latent substance use classes. Bar height represents the prevalence of past-year alcohol use and lifetime use of other substances within each latent class amon participants with non-missing data for the respective indicator. Missing data for each indicator are shown in Table S2. Stacke segments show the distribution of risk categories among participants with use of that substance. Alcohol risk groups were defined using the Alcohol Use Disorders Identification Test–Concise (AUDIT-C) as low risk (0–2 for females and 0–3 for males), moderate risk (3–7 for females and 4-7 for males), and high risk (≥8 for both sexes). Drug use risk was defined using the Alcohol, Smokin and Substance Involvement Screening Test (ASSIST) substance-specific involvement scores as low (0–3), moderate (4–26), and high (≥27); “unknown” indicates lifetime use was reported but the ASSIST involvement score was missing.

### Patterns of polysubstance use in the mixed illicit drug use class

Figure S1 shows the number of participants with each observed combination of lifetime use of cannabis, amphetamines, cocaine, hallucinogens, opioids, and sedatives among participants in the mixed illicit drug use class with complete data on all six substance use indicators (n=46). Overall, 16 participants (34.8%) reported lifetime use of one substance only, 22 (47.8%) reported lifetime use of two substances, and eight (17.4%) reported lifetime use of three or more substances. The most common combination was lifetime use of amphetamines and sedatives (n=14 [30.4%]), whereas other combinations were infrequent.

### Criterion validity of substance use classes

Compared with the low-risk class, the higher-risk alcohol use class was associated with MINI diagnoses of alcohol use disorder (RRR 3.45, 95% CI 2.11–5.64) and substance use disorder (RRR 4.01, 95% CI 1.19–13.46). The cannabis use class was also associated with alcohol use disorder (RRR 2.31, 95% CI 1.27–4.20) and substance use disorder (RRR 5.04, 95% CI 1.42– 17.84). The mixed illicit drug use class was not associated with alcohol use disorder (RRR 1.21, 95% CI 0.38–3.81) but was strongly associated with substance use disorder (RRR 89.11, 95% CI 26.36–301.19) (Figure S2).

### Associations between substance use classes with age, sex, HIV status, and facility

Age and sex were strongly associated with substance use class membership (Figure 2, Figure 3, Figure S3, Figure S4). Compared with participants aged 45 years or older, those aged 18–29 years were less likely to belong to the low-risk class (AME −26.2 percentage points, 95% CI - 35.8 to −16.7) and more likely to belong to the cannabis use class (AME 21.4 percentage points, 95% CI 13.6 to 29.2) (Figure 2). Participants aged 30 to 44 years were also less likely to belong to the low-risk class (AME −20.8 percentage points, 95% CI −30.1 to −11.5) and more likely to belong to the higher-risk alcohol use class (AME 12.7 percentage points, 95% CI 4.3 to 21.2) and mixed illicit drug use class (AME 5.9 percentage points, 95% CI 0.8 to 11.0). Males were less likely to belong to the low-risk class (AME −15.4 percentage points, 95% CI −23.1 to −7.6) and higher-risk alcohol use class (AME −9.6 percentage points, 95% CI −16.7 to −2.5), but more likely to belong to the cannabis use class (AME 22.3 percentage points, 95% CI 15.6 to 29.0). Facility 2 was associated with a substantially higher probability of membership in the mixed illicit drug use class (AME 20.2 percentage points, 95% CI 13.9 to 26.5) and a lower probability of membership in the low-risk class (AME −13.1 percentage points, 95% CI −22.1 to −4.1) (Figure 2). HIV was associated with a 9.0 percentage-point higher probability of membership in the higher-risk alcohol use class (95% CI, 1.0 to 16.9 percentage points; Figure 2), but not with a higher probability of moderate- to high-risk alcohol use (AME, 5.1 percentage points; 95% CI, −3.8 to 14.1; Table S3). The corresponding adjusted predicted probabilities for membership in each substance use class for each level of age group, sex, HIV status, and facility are shown in Figure 3 and Figure S4. For example, the predicted probability of mixed illicit drug use, adjusted for age, sex, and HIV status was 1.4% (95% CI 0.2 to 2.6) in Facility 1 and 21.6% (95% CI 15.5 to 27.7) in Facility 2.

**Figure 2.**
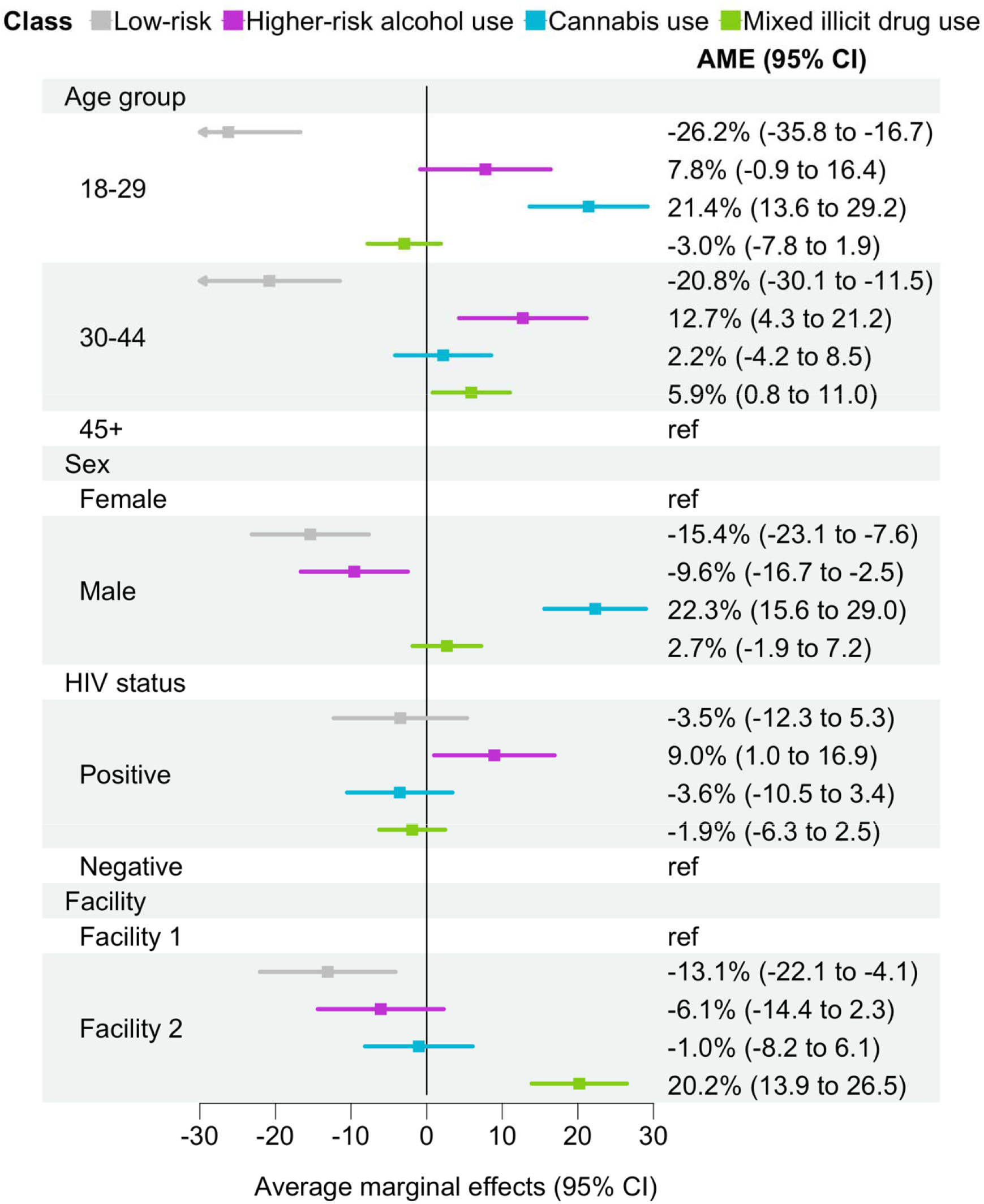
Average marginal effects of age group, sex, HIV status, and facility on substance use clas membership. Adjusted average marginal effects (AME) with 95% confidence intervals (CI) for associations of age group, sex, HIV status, and facility with latent substance use class membership, estimated from multinomial logistic regression. AMEs are expressed as percentage-point differences in the predicted probability of membership in each substance use class, averaging over the observe distribution of the remaining covariates.

**Figure 3.**
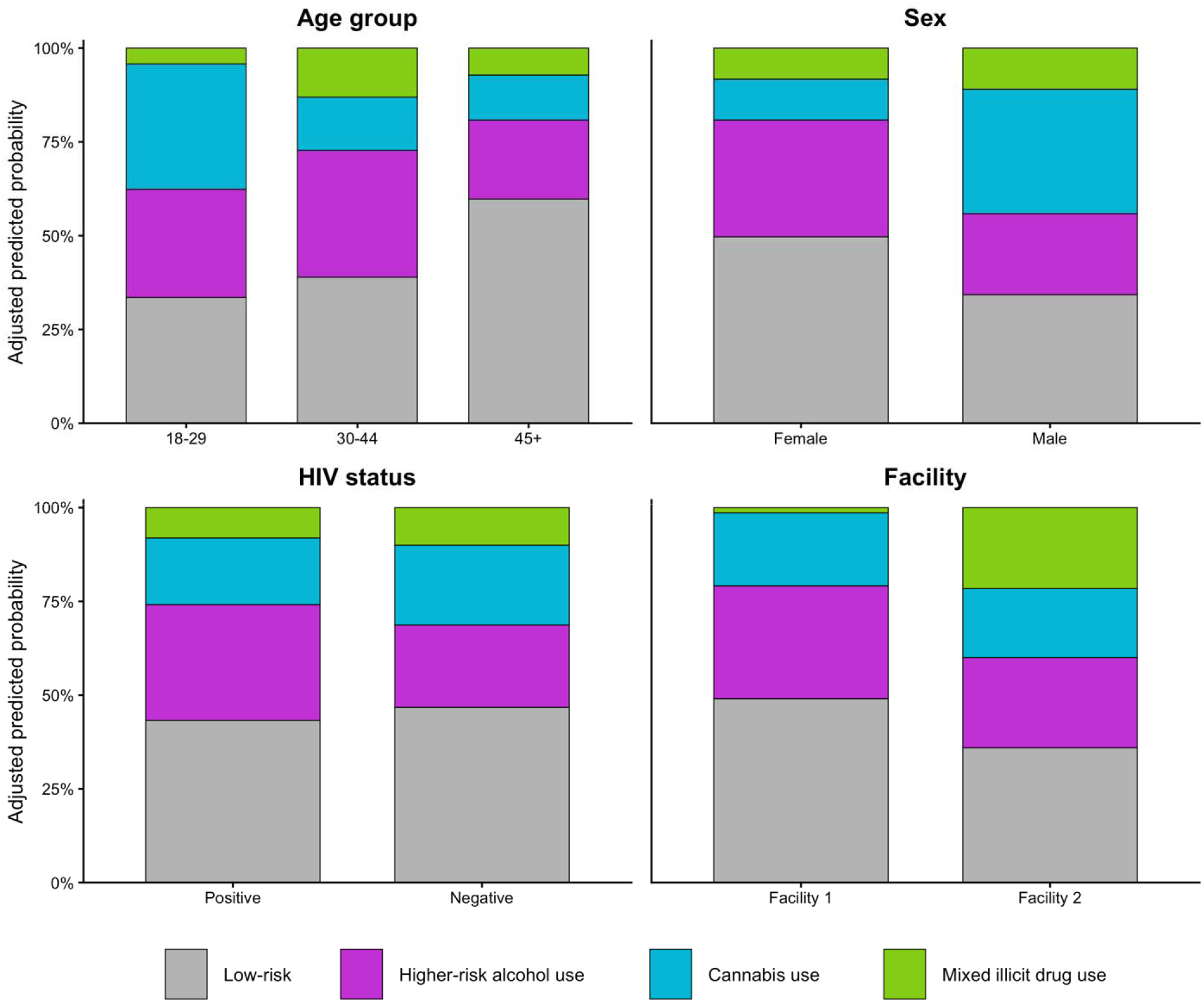
Adjusted predicted probabilities of membership in substance use classes by age group, sex, HIV status, and facility. Stacked bar charts show adjusted predicted probabilities of membership in the four substance use classes by age group, sex, HIV status, and facility. Predicted probabilities were estimated from multinomial logistic regression models adjusting for age group, sex, HIV status, and facility and sum to 100% within each level of a predictor.

## 4. Discussion

In two socioeconomically deprived communities in Cape Town, 44% of participants were classified as low risk, while 56% belonged to higher-risk classes characterized by moderate-to-high-risk alcohol use (28%), cannabis use with mixed alcohol risk (19%), or mixed illicit drug use with little alcohol involvement (9%). Class membership varied substantially by age, sex, and facility, and people with HIV were more likely to belong to the moderate-to-high-risk alcohol class.

The four classes represented qualitatively distinct substance use profiles. The *low-risk class* comprised participants with no or low-risk alcohol use and no lifetime use of other substances. The *moderate-to-high-risk alcohol class* was characterized predominantly by hazardous or harmful alcohol use with relatively limited use of other substances, similar to an alcohol-only class identified among female sex workers with HIV in Durban.^6^ Its clinical relevance was supported by associations with MINI alcohol use disorder, although some participants also met diagnostic criteria for other substance use disorders. The *cannabis class* was characterized by universal cannabis use with heterogeneous levels of alcohol risk, but relatively little use of other illicit substances, consistent with nationally representative and Western Cape treatment data identifying cannabis as a commonly used substance.^3,23^ The cannabis phenotype was associated with both MINI alcohol use disorder and substance use disorder, consistent with the combined alcohol and cannabis use observed. Cannabis-dominant patterns with considerable alcohol use have also been identified in previous South African latent class analyses, including a marijuana-and-alcohol class among women in Cape Town^7^ and a marijuana-and-cigarette class with frequent alcohol use among people who use substances in Pretoria.^8^ The *mixed illicit drug use class* was smaller and qualitatively distinct from both the alcohol- and cannabis-dominant classes. It was characterized by use of multiple substances, particularly amphetamines and sedatives, with limited alcohol use, consistent with Western Cape treatment surveillance, where methamphetamine and methaqualone is commonly used among people seeking substance use treatment, and with population-based estimates showing a smaller subgroup reporting use of multiple illicit drugs.^3,23^ Its clinical distinctiveness was supported by an almost 90-fold higher risk of MINI substance use disorder diagnoses than the low-risk class, with no increase in alcohol use disorder. Previous South African latent class analyses have also identified higher-risk polysubstance classes characterized by combinations of methamphetamine, cannabis, methaqualone, cocaine, heroin, or whoonga, although the specific composition varied across populations. ^6–8^

Age strongly differentiated substance use patterns. Older participants were more likely to be in the low-risk class, whereas younger participants were more often classified in the cannabis-use and moderate-to-high-risk alcohol classes. The observed age pattern is broadly consistent with South African population-based studies showing higher levels of reported substance use and substance use-related admissions among adolescents and younger adults.^3,24^ These age differences may partly reflect greater exposure among younger adults to social and structural conditions associated with substance use, including unemployment, violent victimization, and gang involvement.^5,25^ Mixed illicit drug use was most common at ages 30–44 years. This pattern should be interpreted cautiously, because lifetime measures were used for less commonly used substances dominating the mixed illicit substance use class. Older participants had more time to accumulate lifetime exposure, so differences across age groups reflect differences in exposure opportunity and do not necessarily indicate differences in current use of these substances.

We also observed clear sex differences in substance use patterns. Males were more likely than females to belong to the cannabis-use and mixed illicit drug-use classes, whereas females were more frequently classified in the low-risk class, consistent with South African population-based studies showing higher rates of substance use and substance use-related psychiatric hospital admissions among males.^3,24,25^ Sex differences may reflect gendered social norms around alcohol and substance use,^26,27^ and greater exposure of men to social environments associated with drug use. In South Africa, gang involvement and incarceration are substantially more common among men than among women,^28^ and studies conducted in disadvantaged communities in Cape Town have documented strong associations between gang involvement, contact with the criminal justice system and substance use.^29^ Gang membership often facilitates access to drugs, and involvement in gangs and drug trafficking may be viewed as one of few available ways to generate income in marginalized communities with limited legitimate economic opportunities.^5^ Observed sex differences may also reflect differential reporting. Alcohol and other substance use may be less socially acceptable among females, leading to greater underreporting because of social desirability bias.

Relatedly, we observed pronounced differences between facilities, suggesting that neighborhood characteristics strongly shape local substance use patterns. Mixed illicit drug use was strongly concentrated among participants attending Facility 2, highlighting substantial local heterogeneity within the same metropolitan area. The community served by Facility 2 is located in an area with documented high levels of gang-related violence, indicating substantial gang activity in the surrounding area, which may contribute to local patterns of substance use.^30^ The difference in the prevalence of mixed illicit drug use may reflect neighborhood-specific norms around substance use, including the normalization of methamphetamine use in some communities in Cape Town,^31^ as well as differences in drug availability shaped by local drug markets and gang activity. Such contextual factors can vary substantially between communities in Cape Town and are closely linked to South Africa’s apartheid legacy and persistent spatial inequalities.

HIV status was not consistently associated with a higher prevalence of problematic alcohol or other substance use. Participants with HIV had a significantly higher predicted probability of belonging to the higher-risk alcohol use class by 9.0 percentage points, whereas their probabilities of belonging to the cannabis-use and mixed illicit drug-use classes were lower, but these differences were not statistically significant. However, when individual substances were analyzed separately, HIV status was not associated with moderate-to-high-risk alcohol use or cannabis use and was associated only with a significantly lower probability of amphetamine use. The apparent difference between the class-based and substance-specific findings likely reflects the mutually exclusive nature of the latent classes. Participants with HIV may have been more likely to be classified in the higher-risk alcohol use class because they were less likely to show substance-use patterns characteristic of the cannabis-use or mixed illicit drug-use classes. Importantly, the cannabis-use class also included substantial moderate-to-high-risk alcohol use, so the higher probability of membership in the higher-risk alcohol use class does not necessarily indicate a higher overall prevalence of risky alcohol use among participants with HIV. Overall, our findings do not suggest a greater overall burden of alcohol or substance use among people with HIV, but rather similar levels of risky alcohol use and lower use of some other substances.

The high prevalence of higher-risk alcohol and drug use among adults attending primary care facilities in disadvantaged communities in Cape Town supports the implementation of routine screening and treatment services within these settings. Services should be tailored to heterogeneous local substance use profiles, including prioritizing assessment of locally relevant substances and identifying co-use of alcohol and cannabis or broader patterns of polysubstance use. WHO recommends screening with AUDIT or ASSIST, followed by interventions according to substance-specific risk scores.^20,21,33^ For low-risk use, WHO recommends brief health information, for moderate-risk use a brief intervention in primary care, and for high-risk use or probable dependence more comprehensive assessment and referral to specialist treatment where indicated.

A major strength was the study setting, which enabled us to characterize distinct substance use patterns in socioeconomically disadvantaged peri-urban communities that are not reflected in national estimates. The findings provide policy-relevant evidence to inform screening and treatment services tailored to the needs of marginalized peri-urban populations, which comprise a substantial share of South Africa’s population. We used validated screening tools to measure substance use and MINI diagnostic interviews, providing an external clinical criterion for the identified classes. Latent class analysis further allowed us to identify distinct patterns of concurrent substance use that would be obscured by examining substances separately.

Several limitations should be considered. First, the study included only two primary care communities, limiting our ability to characterize local heterogeneity and potentially missing substance use patterns present in other settings. Second, the use of quota-based sampling and the oversampling of people with HIV limit the generalizability of prevalence estimates to the broader population. Third, some substances were uncommon and therefore combined into broader categories or measured as lifetime use, reducing granularity and comparability across indicators and precluding distinction between experimental and sustained higher-risk use. Fourth, ASSIST and AUDIT-C rely on self-report and may be affected by recall bias and under-reporting. Fifth, ethics guidance restricted reporting of facility names and detailed community characteristics, limiting our ability to interpret heterogeneity of substance use pattern relation to local social and structural context. Finally, the mixed illicit drug-use class was relatively small, resulting in low precision for estimates involving this group.

## Conclusion

Adults attending primary care facilities in socioeconomically disadvantaged peri-urban communities in Cape Town had a high prevalence of higher-risk alcohol and substance use. Substance use was characterized by qualitatively distinct, clinically meaningful profiles, including alcohol-dominant use, cannabis use with heterogeneous levels of alcohol risk, and mixed illicit drug use dominated by amphetamine and sedative use. These profiles differed substantially by age, sex, and study facility, highlighting pronounced demographic and local heterogeneity in substance use patterns. Primary care services in similar settings should integrate routine screening for alcohol and other substance use and tailor assessment and intervention strategies to locally relevant substance use profiles.

## Data Availability

Data were obtained from the International epidemiology Databases to Evaluate AIDS-Southern Africa (IeDEA-SA). Data cannot be made available online because of legal and ethical restrictions. To request data, readers may contact IeDEA-SA for consideration by filling out the online form available at https://www.iedea-sa.org/contact-us/.

## Supplementary appendix

**Table S1.** Fit statistics for latent class models.

| Number of classes | AIC | BIC | Entropy |
| --- | --- | --- | --- |
| 3 | 1571 | 1643 | 0.839 |
| 4 | 1576 | 1671 | 0.779 |
| 2 | 1626 | 1671 | 0.849 |
| 5 | 1585 | 1700 | 0.736 |

Models with 2 to 5 classes were compared and are ordered by ascending BIC. Lower AIC and BIC values indicate better model fit. Entropy ranges from 0 to 1, with higher values indicating clearer class separation of classes. Values greater than 0.80 are commonly interpreted as indicating good separation. AIC=Akaike information criterion. BIC=Bayesian information criterion.

**Table S2.** Alcohol and substance use risk by class.

|  | Low-risk | Higher-risk<br>alcohol use | Cannabis<br>use | Mixed illicit<br>drug use | Total |
| --- | --- | --- | --- | --- | --- |
|  | N=265 (44.0%) | N=168 (27.9%) | N=114 (18.9%) | N=55 (9.1%) | N=602 (100.0%) |
| AUDIT-C risk group, n (%) |  |  |  |  |  |
| Abstinent | 173 (65.3%) | 0 (0.0%) | 18 (15.8%) | 40 (72.7%) | 231 (38.4%) |
| Low | 92 (34.7%) | 0 (0.0%) | 33 (28.9%) | 10 (18.2%) | 135 (22.4%) |
| Moderate | 0 (0.0%) | 135 (80.4%) | 38 (33.3%) | 0 (0.0%) | 173 (28.7%) |
| High | 0 (0.0%) | 23 (13.7%) | 19 (16.7%) | 5 (9.1%) | 47 (7.8%) |
| Missing | 0 (0.0%) | 10 (6.0%) | 6 (5.3%) | 0 (0.0%) | 16 (2.7%) |
| Cannabis risk group, n (%) |  |  |  |  |  |
| None | 265 (100.0%) | 165 (98.2%) | 0 (0.0%) | 40 (72.7%) | 470 (78.1%) |
| Low | 0 (0.0%) | 0 (0.0%) | 42 (36.8%) | 3 (5.5%) | 45 (7.5%) |
| Moderate | 0 (0.0%) | 0 (0.0%) | 65 (57.0%) | 6 (10.9%) | 71 (11.8%) |
| High | 0 (0.0%) | 1 (0.6%) | 4 (3.5%) | 2 (3.6%) | 7 (1.2%) |
| Unknown risk | 0 (0.0%) | 2 (1.2%) | 2 (1.8%) | 0 (0.0%) | 4 (0.7%) |
| Missing | 0 (0.0%) | 0 (0.0%) | 1 (0.9%) | 4 (7.3%) | 5 (0.8%) |
| Amphetamine risk group, n (%) |  |  |  |  |  |
| None | 265 (100.0%) | 161 (95.8%) | 99 (86.8%) | 11 (20.0%) | 536 (89.0%) |
| Low | 0 (0.0%) | 1 (0.6%) | 0 (0.0%) | 8 (14.5%) | 9 (1.5%) |
| Moderate | 0 (0.0%) | 1 (0.6%) | 0 (0.0%) | 19 (34.5%) | 20 (3.3%) |
| High | 0 (0.0%) | 3 (1.8%) | 0 (0.0%) | 12 (21.8%) | 15 (2.5%) |
| Unknown risk | 0 (0.0%) | 2 (1.2%) | 0 (0.0%) | 3 (5.5%) | 5 (0.8%) |
| Missing | 0 (0.0%) | 0 (0.0%) | 15 (13.2%) | 2 (3.6%) | 17 (2.8%) |
| Sedatives risk group, n (%) |  |  |  |  |  |
| None | 265 (100.0%) | 167 (99.4%) | 97 (85.1%) | 28 (50.9%) | 557 (92.5%) |
| Low | 0 (0.0%) | 0 (0.0%) | 1 (0.9%) | 1 (1.8%) | 2 (0.3%) |
| Moderate | 0 (0.0%) | 0 (0.0%) | 1 (0.9%) | 8 (14.5%) | 9 (1.5%) |
| High | 0 (0.0%) | 1 (0.6%) | 0 (0.0%) | 10 (18.2%) | 11 (1.8%) |
| Unknown risk | 0 (0.0%) | 0 (0.0%) | 0 (0.0%) | 3 (5.5%) | 3 (0.5%) |
| Missing | 0 (0.0%) | 0 (0.0%) | 15 (13.2%) | 5 (9.1%) | 20 (3.3%) |
| Cocaine risk group, n (%) |  |  |  |  |  |
| None | 265 (100.0%) | 168 (100.0%) | 93 (81.6%) | 47 (85.5%) | 573 (95.2%) |
| Low | 0 (0.0%) | 0 (0.0%) | 3 (2.6%) | 0 (0.0%) | 3 (0.5%) |
| Moderate | 0 (0.0%) | 0 (0.0%) | 1 (0.9%) | 2 (3.6%) | 3 (0.5%) |
| High | 0 (0.0%) | 0 (0.0%) | 0 (0.0%) | 1 (1.8%) | 1 (0.2%) |
| Unknown risk | 0 (0.0%) | 0 (0.0%) | 3 (2.6%) | 0 (0.0%) | 3 (0.5%) |
| Missing | 0 (0.0%) | 0 (0.0%) | 14 (12.3%) | 5 (9.1%) | 19 (3.2%) |
| Hallucinogens risk group, n (%) |  |  |  |  |  |
| None | 265 (100.0%) | 168 (100.0%) | 98 (86.0%) | 44 (80.0%) | 575 (95.5%) |
| Low | 0 (0.0%) | 0 (0.0%) | 0 (0.0%) | 3 (5.5%) | 3 (0.5%) |
| Moderate | 0 (0.0%) | 0 (0.0%) | 0 (0.0%) | 2 (3.6%) | 2 (0.3%) |
| Unknown risk | 0 (0.0%) | 0 (0.0%) | 1 (0.9%) | 0 (0.0%) | 1 (0.2%) |
| Missing | 0 (0.0%) | 0 (0.0%) | 15 (13.2%) | 6 (10.9%) | 21 (3.5%) |
| Opioids risk group, n (%) |  |  |  |  |  |
| None | 265 (100.0%) | 167 (99.4%) | 96 (84.2%) | 36 (65.5%) | 564 (93.7%) |
| Low | 0 (0.0%) | 0 (0.0%) | 0 (0.0%) | 1 (1.8%) | 1 (0.2%) |
| Moderate | 0 (0.0%) | 0 (0.0%) | 1 (0.9%) | 4 (7.3%) | 5 (0.8%) |
| High | 0 (0.0%) | 0 (0.0%) | 1 (0.9%) | 5 (9.1%) | 6 (1.0%) |
| Unknown risk | 0 (0.0%) | 0 (0.0%) | 0 (0.0%) | 1 (1.8%) | 1 (0.2%) |
| Missing | 0 (0.0%) | 1 (0.6%) | 16 (14.0%) | 8 (14.5%) | 25 (4.2%) |

Data are n (%). Participants with no lifetime drug use and no or low-risk alcohol use, defined as an Alcohol Use Disorders Identification Test-Concise (AUDIT-C) score of 2 or lower for females and 3 or lower for males, were classified a priori into the low-risk class. The remaining three classes were identified using latent class analysis.

**Table S3.** Adjusted marginal effects for associations with moderate- to high-risk alcohol use and lifetime cannabis, amphetamine, or other illicit substance use.

|  | AME, percentage points (95% CI) |  |  |  |
| --- | --- | --- | --- | --- |
|  | Moderate to high-risk alcohol use | Cannabis use | Amphetamine use | Other illicit drug use |
| Age group |  |  |  |  |
| 18-29 | 21.5 (11.9,31.1) | 23.0 (15.1,31.0) | -0.7 (-5.1,3.6) | 1.0 (-4.1,6.0) |
| 30-44 | 17.8 (8.7,26.8) | 4.6 (-2.1,11.2) | 8.2 (3.7,12.8) | 4.7 (0.2,9.1) |
| 45+ | ref | ref | ref | ref |
| Sex |  |  |  |  |
| Female | ref | ref | ref | ref |
| Male | -2.0 (-10.0,6.0) | 24.9 (17.9,31.8) | 2.4 (-1.9,6.7) | 9.2 (4.6,13.8) |
| HIV status |  |  |  |  |
| Positive | 5.1 (-3.8,14.1) | -5.5 (-12.7,1.8) | -5.2 (-9.2,-1.1) | -2.2 (-6.2,1.9) |
| Negative | ref | ref | ref | ref |
| Facility |  |  |  |  |
| Facility 1 | ref | ref | ref | ref |
| Facility 2 | -3.5 (-12.6,5.6) | 2.0 (-5.4,9.5) | 17.4 (11.8,23.0) | 19.2 (12.7,25.7) |

Data are average marginal effects, expressed as percentage-point differences in predicted probability, with 95% confidence intervals for current moderate- to high-risk alcohol use, lifetime cannabis use, lifetime amphetamine use, and lifetime use of other illicit substances, including cocaine, hallucinogens, opioids, and sedatives. Estimates were adjusted for age group, sex, HIV status, and facility. AME, average marginal effect; CI, confidence interval.

**Figure S1.**
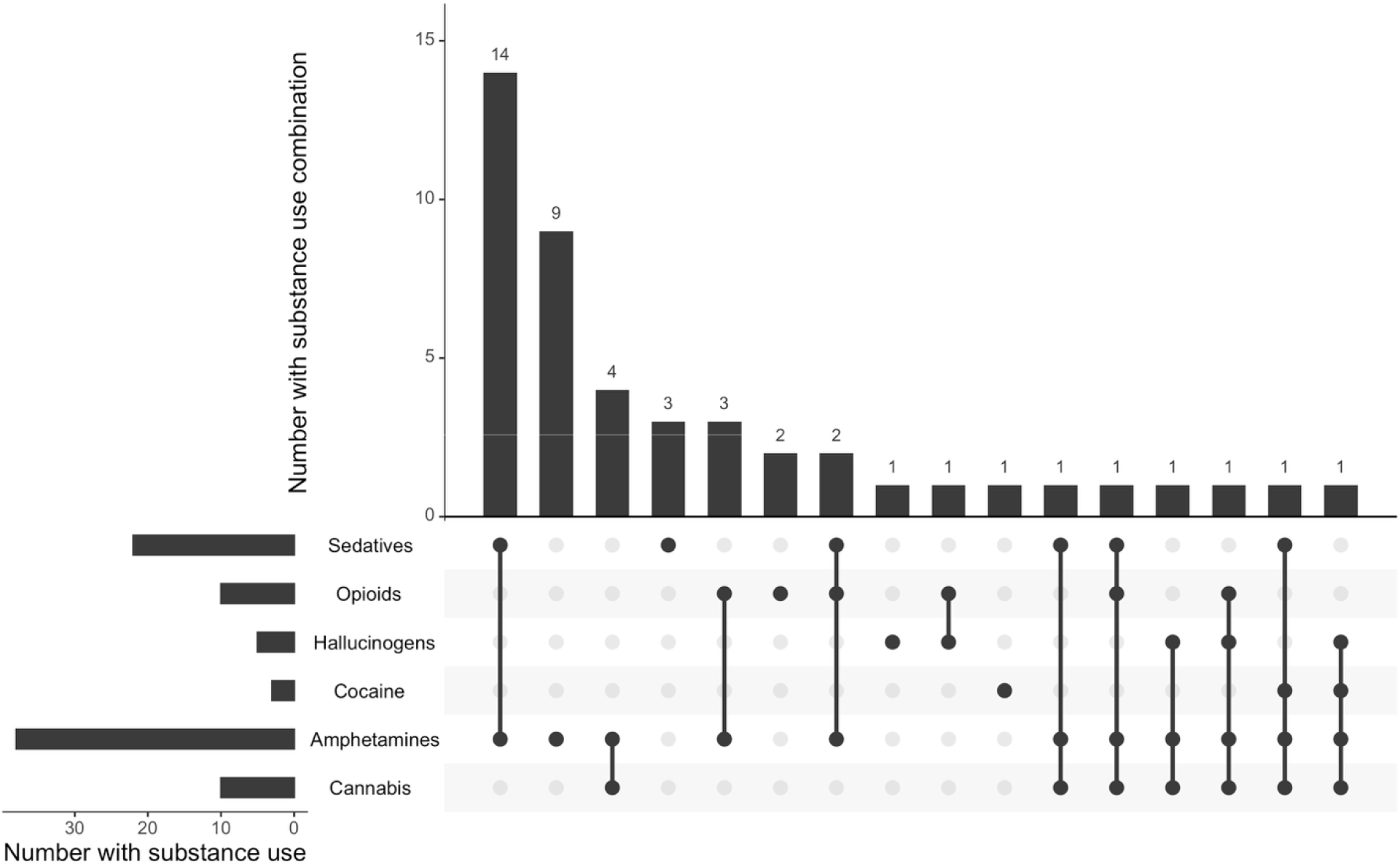
Lifetime substance use combinations in the mixed illicit drug use class. UpSet plot showing combinations of lifetime use of cannabis, amphetamines, cocaine, hallucinogens, opioids, and sedatives amon participants classified in the mixed illicit drug use class. Horizontal bars show the number of participants reporting lifetime use of each substance. Vertical bars show the number of participants with each specific combination. Filled circles indicate substances included in each combination. Only participants with complete data on all six lifetime substance use indicators were included in this analysis (n=46); 9 (16.4%) out of 55 participants in the mixed illicit drug use class were excluded because of missing data.

**Figure S2.**
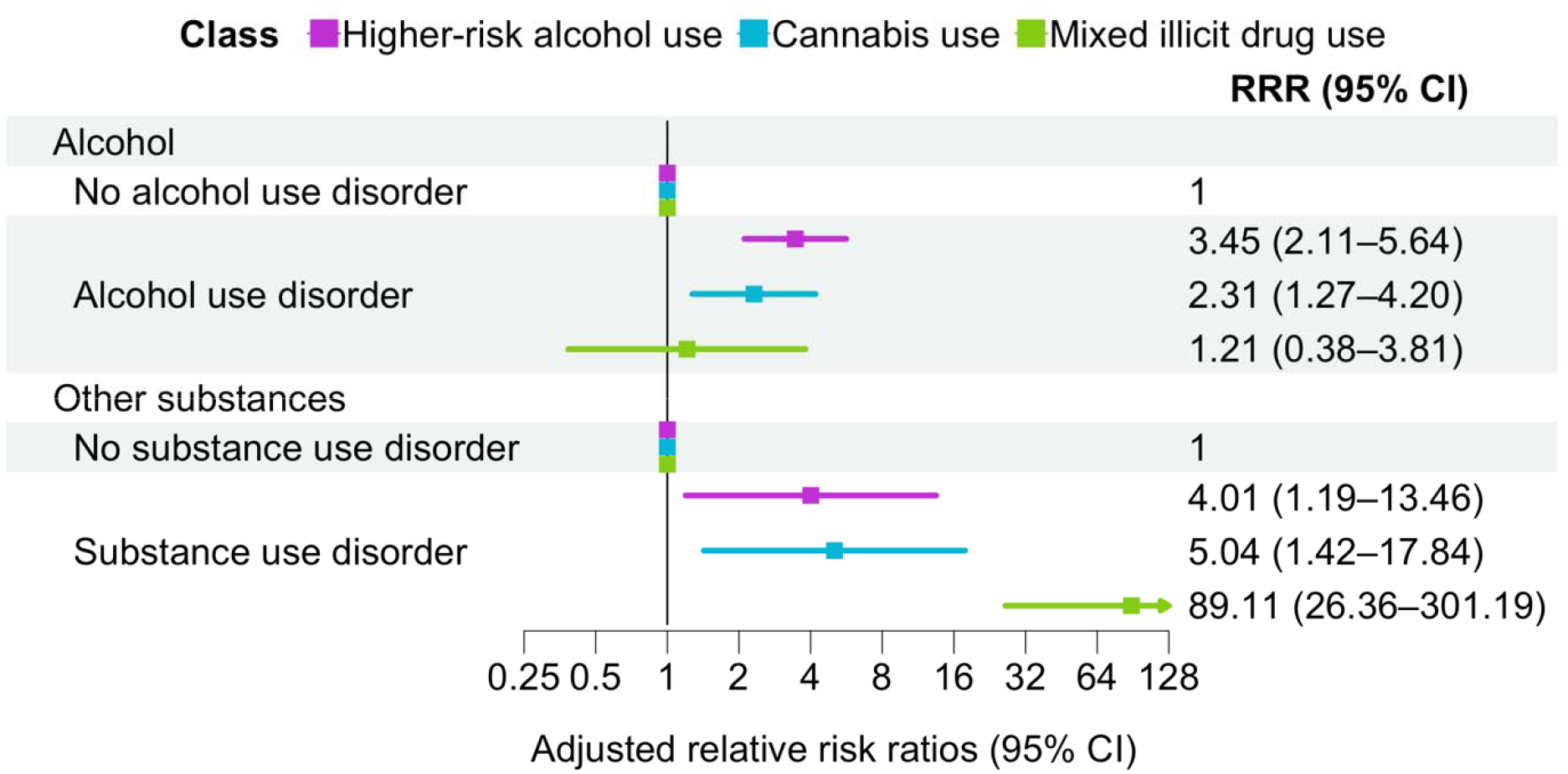
Relative risk ratios for associations between latent substance use classes and MINI diagnoses of alcohol and substance use disorders. Adjusted relative risk ratios (RRR) with 95% confidence intervals (CI) for associations between latent substance use classes and MINI diagnoses of alcohol use disorder and substance use disorder, estimated using multinomial logistic regression with the low-risk class as the reference category. Models were adjusted for age group, sex, HIV status, and facility. MINI, Mini-International Neuropsychiatric Interview.

**Figure S3.**
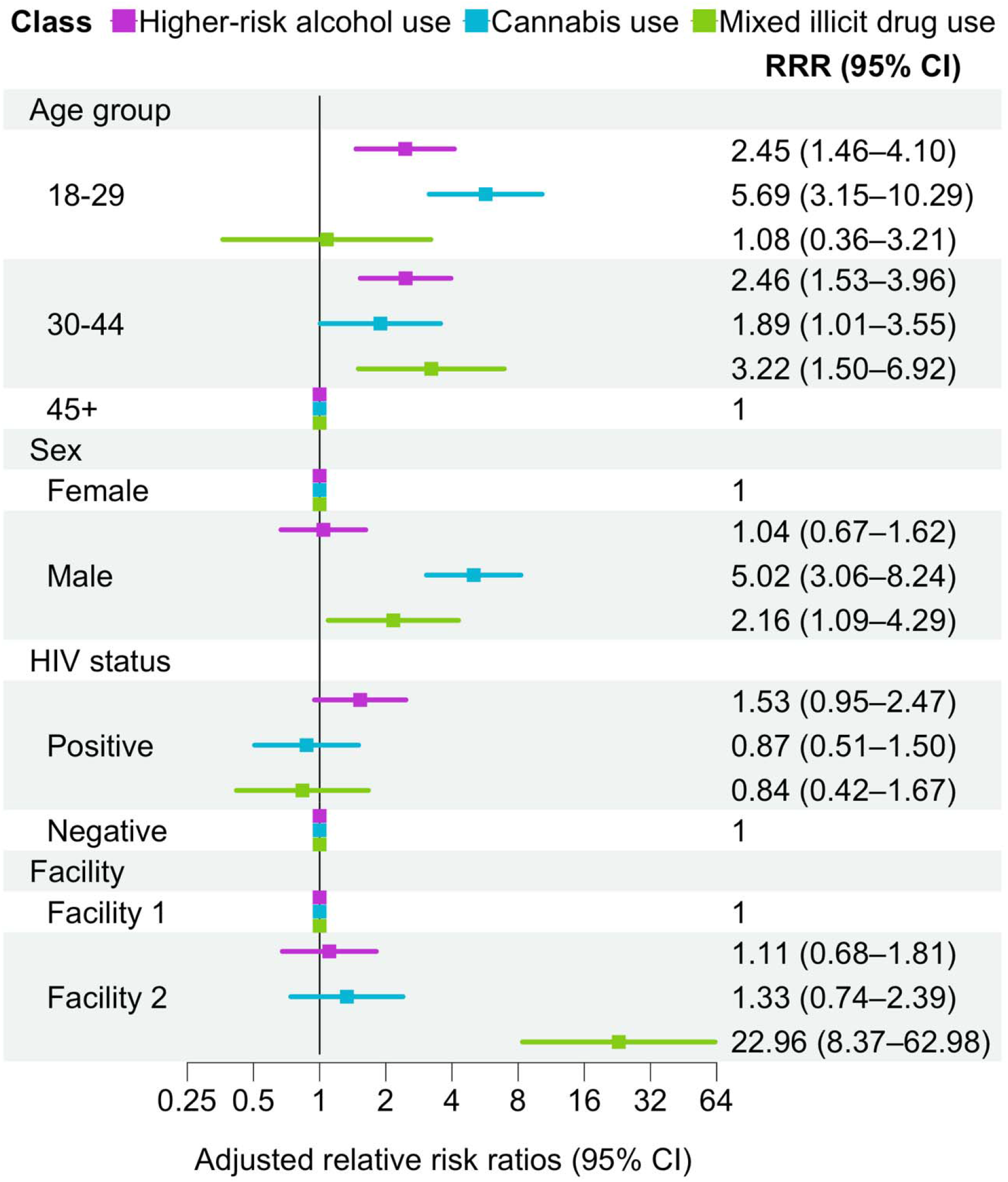
Relative risk ratios for membership in substance use classes compared with the low-risk class. Adjusted relative risk ratios (RRR) with 95% confidence intervals (CI) from multinomial logistic regression for associations of ag group, sex, HIV status, and facility with membership in the higher-risk alcohol use, cannabis use, and mixed illicit drug use classes, compared with the low-risk class.

**Figure S4.**
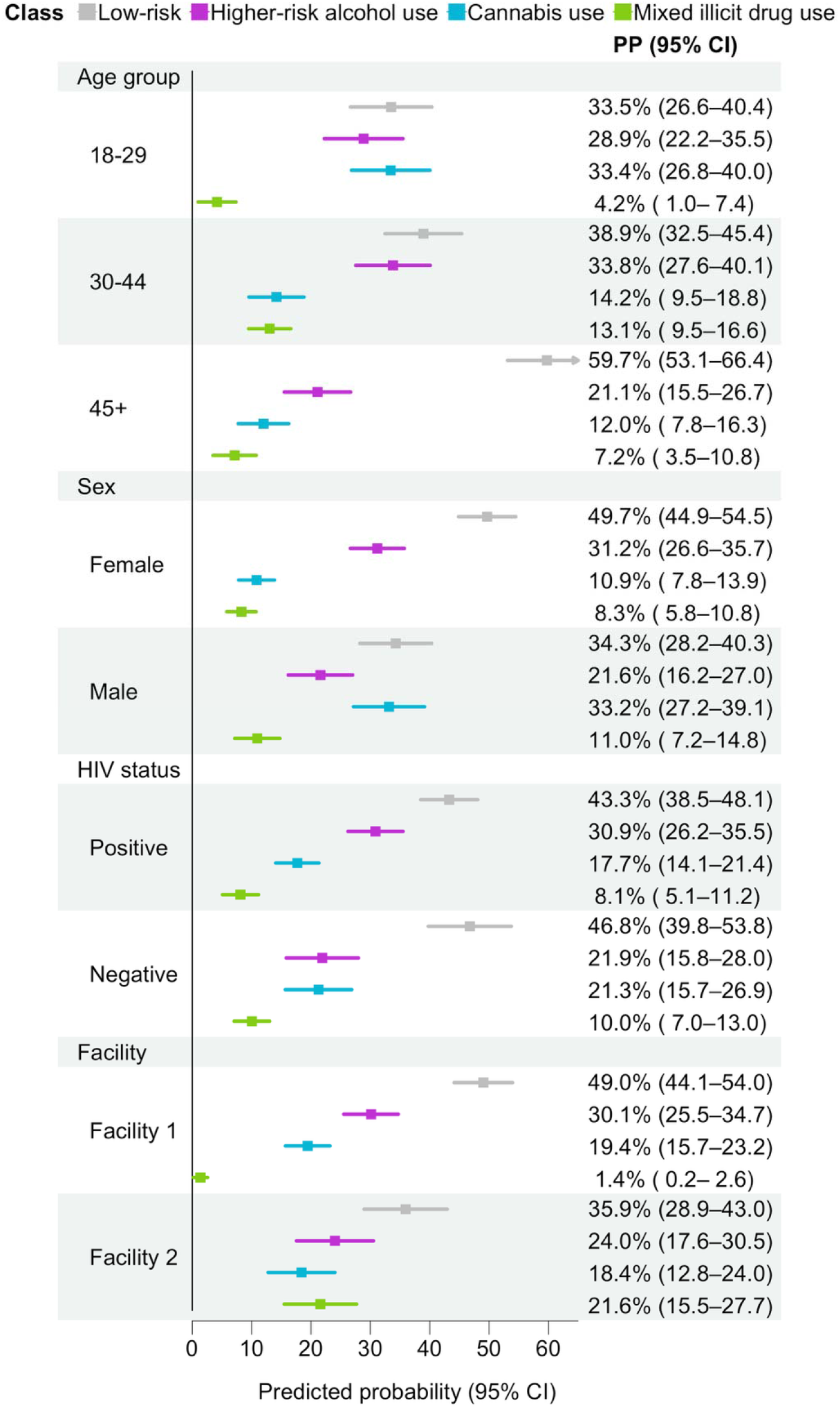
Adjusted predicted probabilities of membership in substance use classes. Adjusted predicted probabilities (PP) with 95% confidence intervals (CI) from multinomial logistic regression for membership in the low-risk, higher-risk alcohol use, cannabis use, and mixed illicit drug use classes by age group, sex, HIV status, and facility, averaging over the observed distribution of the remaining covariates.

